# Active concentration of de novo anti-HLA-DQ donor specific antibodies measured by surface plasmon resonance is associated with chronic lung allograft dysfunction

**DOI:** 10.64898/2026.02.11.26344836

**Authors:** Frédéric Jambon, Carmelo Di Primo, Claire Dromer, Xavier Demant, Antoine Roux, Jerome Le Pavec, Olivier Brugiere, Vincent Bunel, Romain Guillemain, Julien Goret, Marion Duclaut, Marine Cargou, Mamy Ralazamahaleo, Elodie Wojciechowski, Gwendaline Guidicelli, Virginie Hulot, Magali Devriese, Jean-Luc Taupin, Jonathan Visentin

## Abstract

**Background:** In lung transplantation, de novo immunodominant donor-specific anti-HLA antibodies recognizing HLA-DQ antigens (dn-iDSA-DQ) are predominant and can induce chronic lung allograft dysfunction (CLAD). We previously developed a method to measure the active concentration of dn-iDSA-DQ. We aimed to determine whether this new quantitative biomarker is associated with transplantation outcomes.

**Methods:** This retrospective multicentre cohort study included 90 lung transplant recipients (LTRs) developing dn-iDSA-DQ, evidenced through single antigen flow beads (SAFB) follow-up. We measured the active concentration of dn-iDSA-DQ at the time of their first detection (T0) for all LTRs, and within the 2 years after DSA detection, whenever possible. SAFB dn-iDSA-DQ characteristics and clinical data were retrieved up to 5 years after DSA detection.

**Results:** We tested 184 sera with SPR (n=90 at T0, n=94 within the 2 years after DSA detection), among which 63 (34.4%) had a quantifiable concentration of the dn-iDSA-DQ (≥0.3 nM). The median SAFB mean fluorescence intensity (MFI) of the dn-iDSA-DQ with a concentration ≥0.3 nM was higher (p<0.0001), yet the correlation between SAFB MFI and active concentration was low (r=0.758, p<0.0001). In multivariate analysis, a concentration of the dn-iDSA-DQ ≥0.3 nM at T0 was independently associated with a lower 2-year CLAD-free survival (HR 2.06, p=0.02). A concentration of the dn-iDSA-DQ ≥0.3 nM within the 2 years from DSA detection was associated with a lower graft survival in univariate analysis.

**Conclusions:** Active concentration of dn-iDSA-DQ appears as a valuable biomarker to identify pathogenic DSA at their first detection because of its association with CLAD.

## Introduction

Lung transplantation is a treatment option for patients suffering end stage-lung disease which allows to improve quality of life and lung function of selected patients. Several advances have increased lung transplant survival, yet median survival remains limited in comparison with other solid organ transplantations (1). A major cause of graft loss is chronic lung allograft dysfunction (CLAD), defined as the persistent and irreversible decline in graft function (1,2). CLAD is associated with several risk factors, including the development of HLA antibodies directed against the donor (DSA) (3–9).

In lung transplantation, the majority of *de novo* DSA are directed against HLA-DQ and are associated with a worse prognosis (10,11). This knowledge was confirmed thanks to the development of single antigen flow beads (SAFB) assays which greatly improved the resolution and the sensitivity of DSA detection (12–14). However, detection of serum DSA with SAFB has several technical limitations (15–18). Thus, the presence of DSA is not synonymous with lung allograft injury (19) and additional biomarkers are needed to help taking clinical decisions.

SAFB provide a semi-quantitative read-out, the mean fluorescence intensity (MFI), which does not allow to unequivocally identify harmful DSA, although several studies showed an association between DSA MFI level and clinical outcomes (11,20,21). Some authors have also observed worse clinical outcomes in case of multiple DSA (22). Another information available with classical SAFB assays along patient follow-up is the persistence of DSA, which is of worse prognosis in numerous studies (13,22–25).

In parallel, the ability of DSA to fix (C1q SAFB assay) or activate (C3d SAFB assay) complement on SAFB was associated with CLAD and graft loss (20,22,26) and has been proposed to evaluate DSA functionality *in vivo*. However, in C1q and C3d SAFB assays, the ability of DSA to activate complement *in vitro* is strongly associated with SAFB MFI. This means that these modified SAFB assays are not able to detect all complement-activating DSA which are the most prevalent as evidenced by DSA subclass analyses (27–29). Finally, analysing DSA eluted from graft biopsies with SAFB is another mean to evidence DSA that do interact with the graft yet it requires an invasive procedure (30–32).

We hypothesized that the determination of truly quantitative parameters of the DSA, for instance their active concentration, *i.e.* the fraction that really binds to the antigen, and their affinity for the donor antigen, would help understanding their pathogenic potential. We developed a method using surface plasmon resonance (SPR) allowing an accurate measurement of the active concentration and the affinity of serum anti-HLA antibodies (33–35). This includes anti-HLA-DQ which are very often the immunodominant DSA, *i.e.* the one with the highest SAFB MFI, in recipients having more than one DSA. In this retrospective multicentre study, our main goal was to evaluate whether the active concentration of de novo immunodominant anti-DQ DSA (dn-iDSA-DQ) at the time of their first detection was a potential new biomarker in lung transplantation.

## Materials and methods

### Patient selection

This retrospective multicentre study included lung transplant recipients (LTRs) from five transplant centres: Bordeaux University Hospital, Foch Hospital, Marie Lannelongue Hospital, Bichat Hospital and Georges Pompidou Hospital (NCT03474536). Among the LTRs transplanted from January 2001 to December 2021 who had a regular follow-up with SAFB, we identified 211 LTRs who developed dn-iDSA-DQ (MFI threshold of 500). We did not include the LTRs having iDSA which we were not able to study by SPR because of a competition with the capture anchor (see below and Supplemental Figure 1, n = 47), or because of an insufficient quantity of serum in laboratory’s biobanks (Supplemental Figure 1, n = 62). Eight more patients had exclusion criteria (age, study refusal, preformed DSA), leading to 94 LTRs included. Among those, four were excluded because of early infection-related death 1 to 4 months after dnDSA development (n=3) or because of a restrictive allograft syndrome (RAS) related to relapse of the initial disease (n=1).

### Clinical outcomes

We collected clinical data from the transplantation to the post-transplant period up to five years after dnDSA development. The study was approved by the institutional review board and did not interfere with standard patient clinical management. All but one LTRs received conventional triple-drug immunosuppression associating methylprednisolone, a calcineurin inhibitor (cyclosporine A or tacrolimus) and a cytostatic agent (azathioprine or mycophenolate mofetil). The definition of CLAD was based on the International Society for Heart and Lung Transplantation 2019 Consensus criteria, namely a persistent decline (>20%) measured FEV1 value from the reference (baseline) value (36). Baseline transplant function was established as the mean of the best 2 FEV1 measurements taken >3 weeks apart. CLAD was calculated in an automated fashion, and each case was verified by a physician review of medical records. CLAD was further phenotyped as restrictive allograft syndrome (RAS), when appropriate. Graft loss was defined as recipient’s death due to graft dysfunction, replacement on the waiting list or re-transplantation. Antibody-mediated rejection (AMR) was defined according to the consensus report of the International Society for Heart and Lung Transplantation (37).

### Serum selection

We studied with SPR the first serum which evidenced the dn-iDSA anti-DQ for the large majority of LTRs included (86.7%, see results section). Whenever possible, we tested additional sera obtained between 6 and 12 months (T1) and/or between 12 and 24 months (T2) after DSA detection.

### DSA – Anchor competition experiments

Because the size of the anchor used in SPR experiments is the same as the size of the DSA tested, we anticipated that our experiments could suffer from competition between the anchor and the DSA. Thus, we tested for competition between dn-iDSA-DQ and the anchor used in SPR experiments before patient inclusion. Briefly, 17 µl of raw or diluted patients’ sera were EDTA-treated (2 µl) to avoid complement interference (17,38) and were mixed with 1 µl of PBS or 1 µl of the SPR anchor (anti-DQ antibody Tu169, BD Biosciences, 25 µg/ml final concentration). These two mixes were then tested with SAFB following manufacturer’s recommendations (LabScreen LS2A01 SAFB assay, One Lambda, Thermofisher) and analysed on a Luminex 100^®^ analyser (Luminex, Austin, TX). We considered that there was a competition between the DSA and the anchor for a ratio (DSA MFI with anchor) / (DSA MFI with PBS) below 0.7, based on SAFB interassay reproducibility (Supplemental Figure 2), which represented 47 sera (22.3%).

### Serum IgG purification

Before studying sera with SPR, we purified serum IgG with G protein using spin columns from ThermoFisher Scientific, as previously described (34). After protein G purification, samples were concentrated with 10 kDa cut-off Amicon Ultra centrifugal units (Merck-Millipore, Molsheim, France). Total IgG concentrations in serum samples (initial IgG concentration) and purified IgG samples (final IgG concentration) were measured by nephelometric method on a BNII system (Siemens Healthcare Diagnostics, Marburg, Germany) with N antiserum to human IgG (Siemens Healthcare Diagnostics). As the purification process involved IgG constant fragments binding and not IgG variable fragments, we assumed that IgG anti-HLA antibodies present in serum behaved similarly to the other serum IgG. Then, the active concentration of anti-HLA antibodies initially present in the serum was obtained by multiplying the active concentration measured in the purified IgG sample by the ratio of initial total IgG concentration/final total IgG concentration.

### Active concentration measurement

SPR experiments were performed as described previously (33–35). We injected the purified IgG containing the DSA over the surface covered with the HLA-DQ molecule antigens which they recognized, and we did the same with a control HLA-DQ antigen, chosen from the recipient HLA DNA typing and serum SAFB antibody analysis. The lower limit of quantification of 0.3 nM was determined previously (34). The full workflow for active concentration determination is shown in Supplemental Figure 3.

### Statistical analysis

The independent-samples Mann–Whitney U-test and the Fisher’s exact test were used for group comparisons of continuous and categorical variables, respectively. Pearson correlation coefficient was used to correlate quantitative variables. Kaplan–Meier analysis was used to construct graft survival curves. Comparisons used the log-rank test. A Cox proportional hazards analysis was used for univariate and multivariate analyses for CLAD-free or graft survival. Multivariate analyses included variables that showed trends in univariate analysis, i.e. p ≤ 0.2. Variables not independently predictive of graft survival were dropped using the backward elimination procedure. Results were reported as hazard risk (HR) with a 95% confidence interval (CI) and corresponding p-value. P-value ≤ 0.05 was considered statistically significant. Analyses were performed with MedCalc software (Mariakerke, Belgium).

## Results

### Patient characteristics at transplantation and at the time of dn-iDSA-DQ detection

Table 1 shows LTRs characteristics at transplantation. As defined in our inclusion criteria, none of them had preformed DSA. Supplemental Table 1 shows clinical events which occurred between transplant and dnDSA detection. Recipients were followed up to 60 months post dn-iDSA-DQ development, with a mean of 41 months. Dn-iDSA-DQ were mainly detected in the first-year post-transplantation (median 3 months, 1^st^ and 3^rd^ quartiles 1 – 10 months). We studied with CFCA in SPR the first serum in which the dn-iDSA-DQ were detected with SAFB for the large majority of LTRs included (n=78, 86.7%). For the 12 remaining LTRs, the median delay between the first serum in which the dn-iDSA-DQ were detected with SAFB and the serum tested by SPR was 0.7 month (1^st^-3^rd^ quartiles, 0.3 - 0.8 months). Moreover, the median delay between the first serum tested by SPR and the last DSA-negative serum was 2.3 months (10^th^-90^th^ percentiles, 0 – 9.5 months, range 0.2 – 18.7 months). Altogether, these data indicate that we were able to study dn-iDSA-DQ concentration as early as possible in the natural history of allogeneic humoral response of these LTRs.

**Table 1.** Lung recipients’ characteristics at transplant. COPD: chronic obstructive pulmonary disease; PAH: pulmonary arterial hypertension; MMF: mycophenolate mofetil.

|  |  | n = 90 recipients |
| --- | --- | --- |
| Recipient gender (F/M) |  | 34/56 |
| Recipient age [median (1Q/3Q), years] |  | 50 (38 - 60) |
| Initial disease (n, %) |  |  |
|  | Cystic fibrosis | 16 (17.8%) |
|  | COPD/Emphysema | 32 (35.6%) |
|  | Pulmonary fibrosis | 13 (14.4%) |
|  | PAH | 17 (18.9%) |
|  | Others | 12 (13.3%) |
| Pre-transplant allogeneic events (n, %) |  |  |
|  | Transfusion | 2 (2.2%) |
|  | Pregnancy | 11 (12.2%) |
|  | Previous transplantation | 2 (2.2%) |
| Donor gender (F/M) |  | 30/60 |
| Donor age [median (1Q/3Q)] |  | 49 (31 - 63) |
| Donor smoking (n, %) |  | 29 (32.2%) |
| Donor pneumopathy (n, %) |  | 14 (15.6%) |
| CMV status (n, %) |  |  |
|  | D+/R+ | 21 (23.3%) |
|  | D+/R- | 19 (21.1%) |
|  | D-/R- | 30 (33.3%) |
|  | D-/R+ | 20 (22.2%) |
| Transplantation type (n, %) |  |  |
|  | Double-lung | 80 (88.9%) |
|  | Single-lung | 6 (6.7%) |
|  | Heart-lung | 4 (4.4%) |
| Cold ischemia (minutes) |  |  |
|  | Right [median (1Q/3Q)] | 288 (240 - 346) |
|  | Left [median (1Q/3Q)] | 356 (287 - 385) |
| Lung reduction (n, %) |  | 10 (11.1%) |
| Post-operative ECMO (n, %) |  | 12 (13.3%) |
| Primary graft failure (n, %) |  | 10 (11.1%) |
| Induction therapy (anti-IL2R/ATG/none) (n, %) |  | 4 (4.44%) /31 (34.4%) /54 (60%) |
| Initial maintenance therapy |  |  |
|  | Ciclosporin/Tacrolimus (n, %) | 75 (83.3%) /15 (16.7%) |
|  | Steroids (n, %) | 89 (98.9%) /1 (1.1%) |
|  | MMF/Azathioprine (n, %) | 89 (98.9%) /1 (1.1%) |

### Dn-iDSA-DQ characteristics

At T0 (first serum tested with SPR), the median SAFB MFI of the dn-iDSA-DQ was 4469 (1^st^ and 3^rd^ quartiles 2224-9693, range 528-23155). The dn-iDSA-DQ was associated with one or more additional DSA in 53 (58.9%) LTRs. In order to study the evolution of DSA active concentration and its association with clinical outcomes, we also measured dn-iDSA-DQ concentration in one or two additional at T1 (6-12 months after DSA development) and/or T2 (12 – 24 months after DSA development) for 22 (24.4%) and 36 (40%) LTRs had the corresponding serum available. Altogether, we then tested 184 sera with SPR, among which 63 (34.4%) showed an active concentration of the dn-iDSA-DQ ≥ 0.3 nM (quantification threshold, see Materials and methods section). The SAFB MFI of the dn-iDSA-DQ with an active concentration ≥ 0.3 nM was higher than those which were not quantifiable (Mann-Whitney U-Test, p < 0.0001, Figure 1A). Taking into consideration all the tested sera, there was a low correlation between SAFB MFI and active concentration (r = 0.758, 95% CI 0.688 – 0.813, after logarithmic transformation of active concentration Figure 1B). We obtained similar results when analysing the 63 sera showing an active concentration of the dn-iDSA-DQ ≥ 0.3 nM only (r = 0.707, 95% CI 0.556 – 0.812, after logarithmic transformation of active concentration). Seeking for more information about the relation between DSA characteristics, we observed that all the 30 dn-iDSA-DQ with an active concentration ≥ 0.3 nM at T0 persisted over time according to SAFB results retrieved from the medical and laboratory files. This was the case for 25 among the 59 (42.4%) which were not quantifiable (Fisher’s exact test p < 0.000001, 1 patient among the 90 was excluded from the analysis due to early death). Similarly, 34 (77.3%) of the dn-iDSA-DQ with a T0 MFI above the median (MFI > 4469) persisted over time, whereas this was the case for only 21 among the 45 (46.7%) with a T0 MFI below the median (Fisher’s exact test p = 0.004).

**Figure 1:**
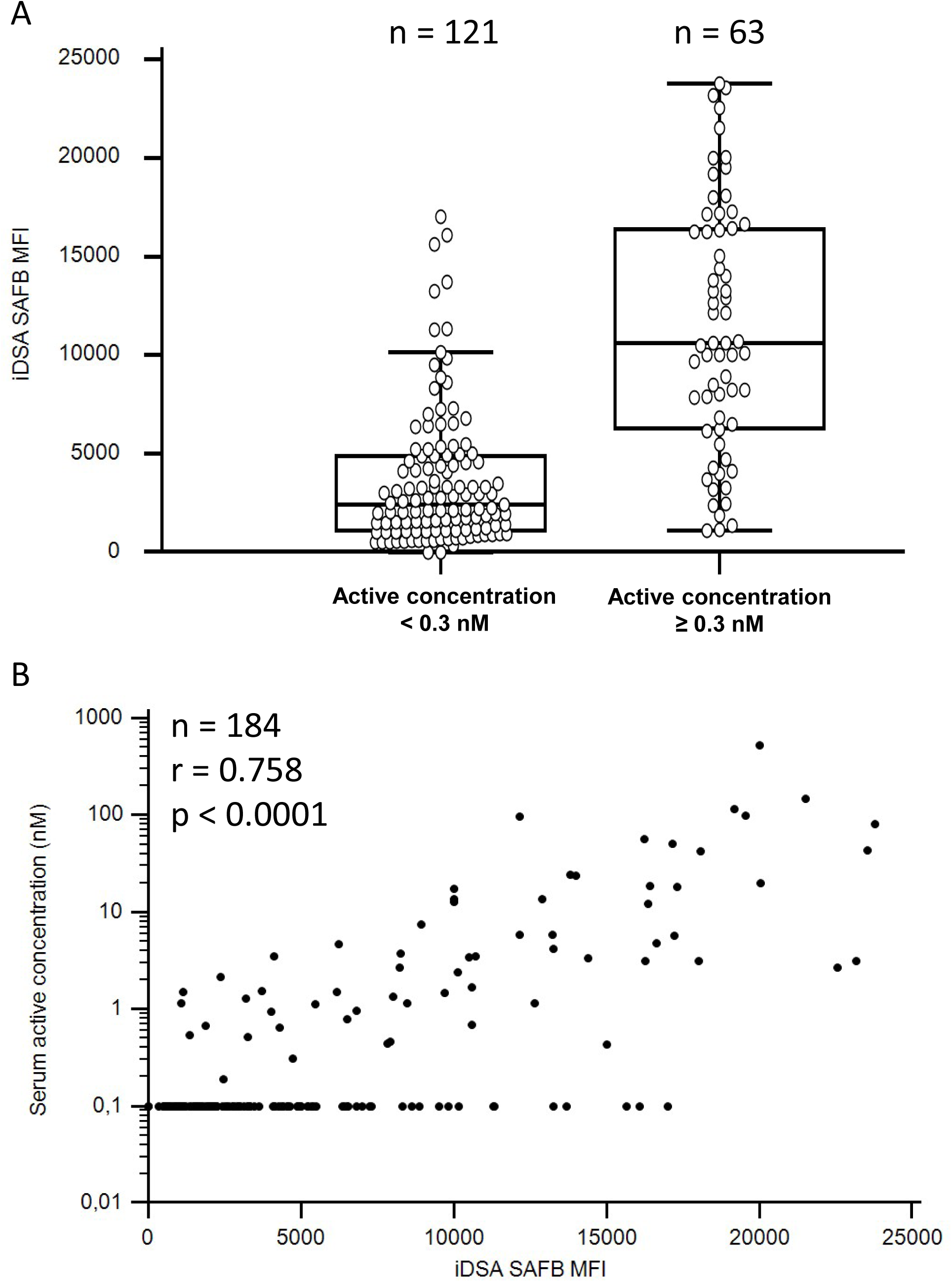
Relation between SAFB MFI and active concentration of dn-iDSA-DQ. (A): Comparison of SAFB MFI between dn-iDSA-DQ with an active concentration below or above the quantification threshold (0.3 nM) (Mann-Whitney U-test). (B): Correlation between SAFB MFI and active concentration of dn-iDSA-DQ (Pearson coefficient correlation). dn-iDSA-DQ: de novo immunodominant donor-specific anti-HLA-DQ antibodies; MFI: mean fluorescence intensity; SAFB: single antigen flow beads.

### An active concentration of dn-iDSA-DQ ≥ 0.3 nM at their first detection is associated with lower CLAD-free survival

The primary objective of this study was to determine whether the active concentration of dn-iDSA-DQ at the time of their first detection with SAFB was associated with graft dysfunction within the 2 years after DSA development. Forty-eight (53.9%) of the 90 LTRs developed CLAD within the 2 years after DSA development. Only 8 LTRs had an AMR diagnosed together with dn-iDSA-DQ detection. Thirty-one (34.4%) had an active concentration of their dn-iDSA-DQ ≥ 0.3 nM. LTRs having an active concentration of their dn-iDSA-DQ above this threshold had a lower CLAD-free survival (Figure 2A, log-rank test, p = 0.02). Regarding SAFB parameters, the dn-iDSA-DQ SAFB MFI was not associated with CLAD-free survival when comparing those below and those above the median MFI (MFI > 4469) (Figure 2B, log-rank test, p = 0.10). We observed the same trend using MFI as a continuous variable with a univariate Cox hazard regression analysis. The LTRs having multiple DSA had a lower CLAD-free survival (Figure 2C, log-rank test, p = 0.002). Several other clinical events occurring between the transplant and dnDSA detection were associated with CLAD-free survival in univariate analyses, *i.e.* biopsy-proven ACR and different types of pulmonary infections (Table 2). Multivariate analysis identified several factors independently associated with lower CLAD-free survival: dn-iDSA-DQ active concentration ≥ 0.3 nM (HR 2.06, 95% CI 1.14 – 3.73, p = 0.02), multiple DSA (HR 3.07, 95% CI 1.61 – 5.84, p = 0.0007), biopsy-proven acute cellular rejection (HR 2.45, 95% CI 1.37 – 4.39, p = 0.003) and bacterial pulmonary infection (HR 2.80, 95% CI 1.24 – 6.33, p = 0.01). We repeated the analyses with a follow-up lasting up to 5 years after dn-iDSA-DQ detection and we observed similar results (Supplemental Table 2).

**Figure 2:**
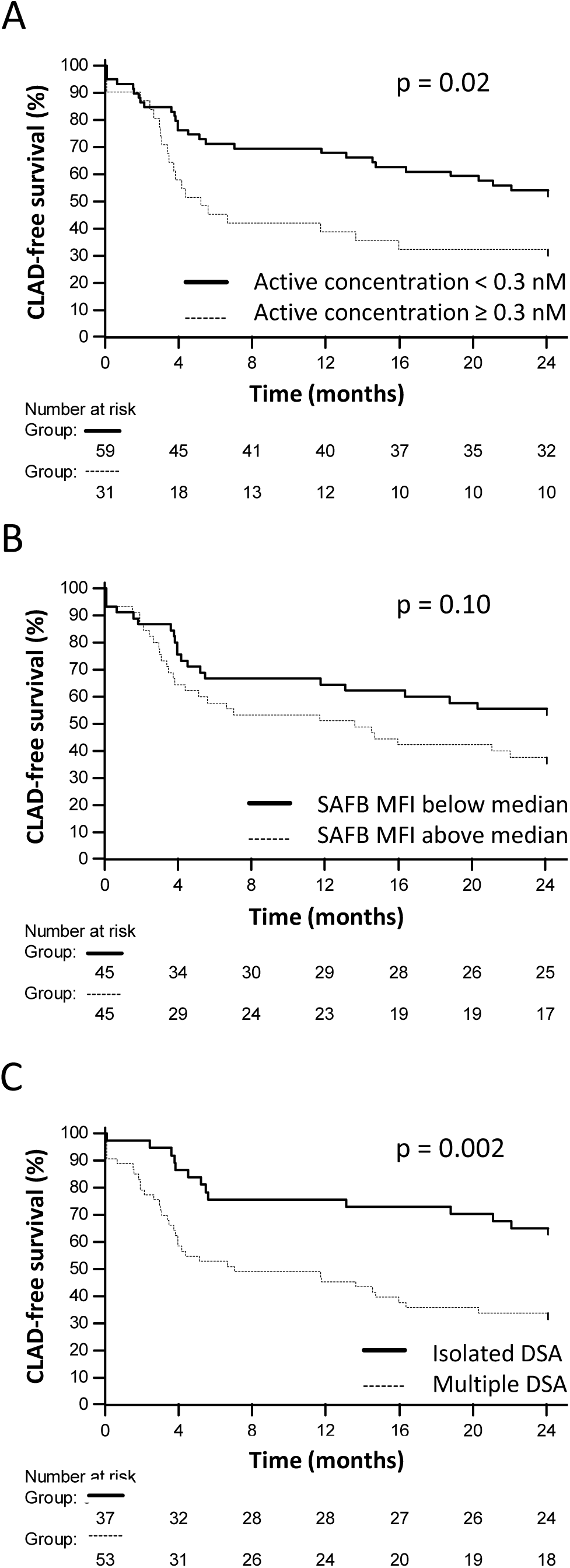
Association between dn-iDSA-DQ characteristics at their first detection and 2-year CLAD-free survival. Comparison of CLAD-free survival between: (A) recipients having an active concentration of their dn-iDSA-DQ below or above 0.3 nM at their first detection; (B) recipients having a SAFB MFI below or above the median at their first detection, (C): recipients having isolated or multiple DSA at their first detection. CLAD: chronic lung allograft dysfunction; dn-iDSA-DQ: de novo immunodominant donor-specific anti-HLA-DQ antibodies; MFI: mean fluorescence intensity; SAFB: single antigen flow beads.

**Table 2:**
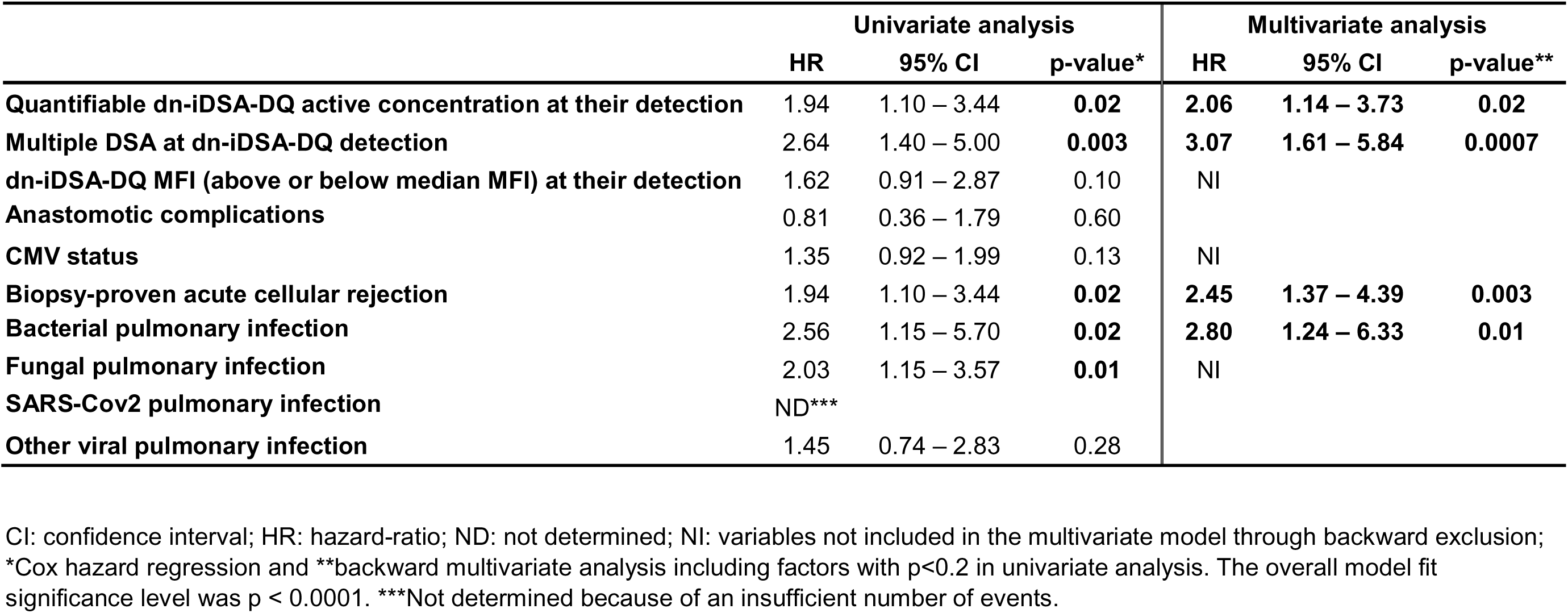
Factors from transplantation to dn-iDSA-DQ detection associated with CLAD at 2 years after DSA detection.

|  | Univariate analysis |  |  | Multivariate analysis |  |  |
| --- | --- | --- | --- | --- | --- | --- |
|  | HR | 95% CI | p-value* | HR | 95% CI | p-value** |
| **Quantifiable dn-iDSA-DQ active concentration at their detection** | 1.94 | 1.10 – 3.44 | **0.02** | **2.06** | **1.14 – 3.73** | **0.02** |
| **Multiple DSA at dn-iDSA-DQ detection** | 2.64 | 1.40 – 5.00 | **0.003** | **3.07** | **1.61 – 5.84** | **0.0007** |
| **dn-iDSA-DQ MFI (above or below median MFI) at their detection** | 1.62 | 0.91 – 2.87 | 0.10 | NI |  |  |
| **Anastomotic complications** | 0.81 | 0.36 – 1.79 | 0.60 |  |  |  |
| **CMV status** | 1.35 | 0.92 – 1.99 | 0.13 | NI |  |  |
| **Biopsy-proven acute cellular rejection** | 1.94 | 1.10 – 3.44 | **0.02** | **2.45** | **1.37 – 4.39** | **0.003** |
| **Bacterial pulmonary infection** | 2.56 | 1.15 – 5.70 | **0.02** | **2.80** | **1.24 – 6.33** | **0.01** |
| **Fungal pulmonary infection** | 2.03 | 1.15 – 3.57 | **0.01** | NI |  |  |
| **SARS-Cov2 pulmonary infection** | ND*** |  |  |  |  |  |
| **Other viral pulmonary infection** | 1.45 | 0.74 – 2.83 | 0.28 |  |  |  |

CI: confidence interval; HR: hazard-ratio; ND: not determined; NI: variables not included in the multivariate model through backward exclusion;
\*Cox hazard regression and \*\*backward multivariate analysis including factors with $p < 0.2$ in univariate analysis. The overall model fit significance level was $p < 0.0001$ . \*\*\*Not determined because of an insufficient number of events.

### An active concentration of dn-iDSA-DQ ≥ 0.3 nM within the 2 years from their detection is associated with graft loss

We then sought to determine whether additional DSA characteristics available within the 2 years from DSA detection, including the day of their detection, were associated with graft loss. The sera studied with SPR along the 2 years from dn-iDSA-DQ detection identified 5 LTRs having an active concentration < 0.3 nM at T0 but ≥ 0.3 nM at T1 or T2. Moreover, 55 (61.1%) LTRs had persistent dn-iDSA-DQ with SAFB assays within 6 to 24 months after DSA detection. LTRs having an active concentration of their dn-iDSA-DQ ≥ 0.3 nM at T0, T1 and/or T2 also had a lower graft survival in univariate analysis (Figure 3A, log-rank test, p = 0.02), which was not the case of those having multiple DSA (log-rank test, p = 0.09, data not shown). However, none of the LTRs having an isolated dn-iDSA-DQ with an active concentration < 0.3 nM lost their graft (Figure 3B, log-rank test, p = 0.007). LTRs having persistent DSA with SAFB also had a lower graft survival in univariate analysis (Figure 3C, log-rank test, p = 0.02).

**Figure 3:**
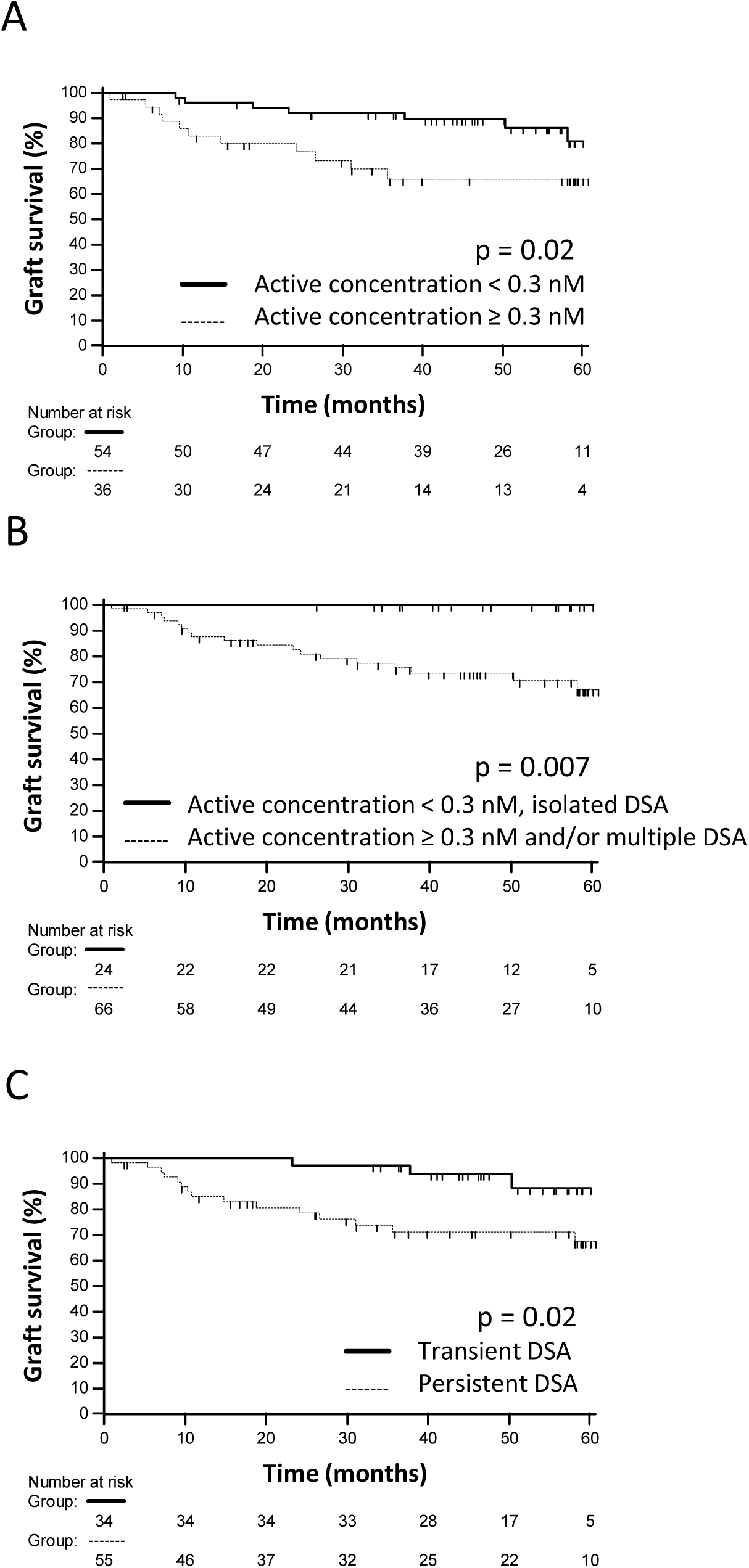
Association between dn-iDSA-DQ characteristics within 2-years from their detection and 5-year graft-survival. Comparison of graft survival between: (A) recipients having an active concentration of their dn-iDSA-DQ below or above 0.3 nM at their first detection and/or within the 2 years after their detection; (B) recipients having an isolated dn-iDSA-DQ with an active concentration < 0.3 nM and recipients having multiple DSA and/or a dn-iDSA-DQ with an active concentration ≥ 0.3; (C) recipients having transient or persistent DSA with SAFB. dn-iDSA-DQ: de novo immunodominant donor-specific anti-HLA-DQ antibodies; SAFB: single antigen flow beads.

In parallel, AMR was diagnosed over the 2 years post-dn-iDSA-DQ detection in 46 (51.1%) LTRs. An active concentration ≥ 0.3 nM of dn-iDSA-DQ at T0, T1 and/or T2, SAFB MFI of dn-iDSA-DQ above the median at T0, multiple DSA at T0 or DSA persistence were not associated with AMR (Supplemental table 3). However, AMR alone, or combined with these four DSA characteristics were associated with graft loss in univariate analysis (Figure 4).

**Figure 4:**
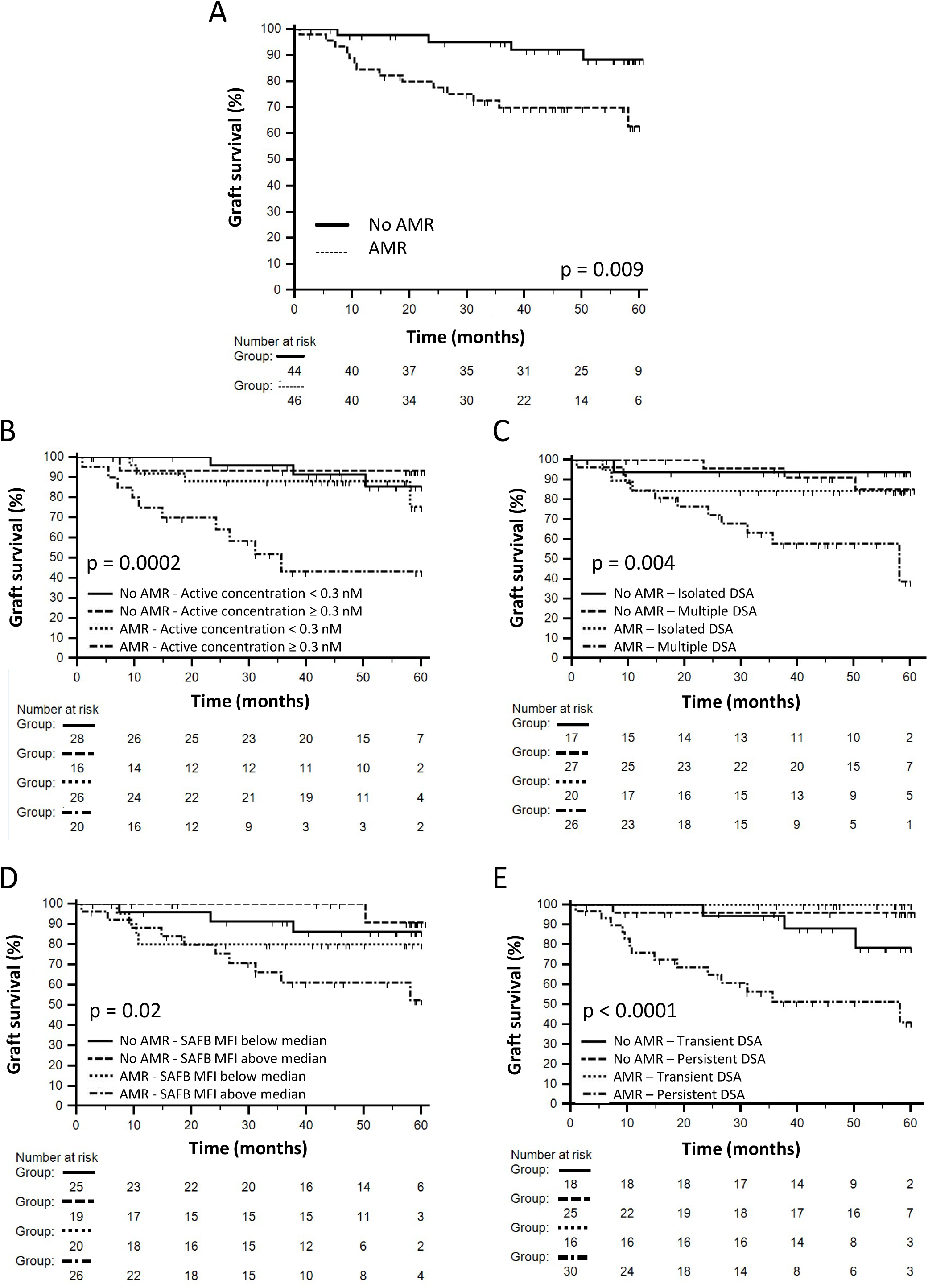
Association between AMR and dn-iDSA-DQ characteristics with 5-year graft-survival. (A) Comparison of graft survival between recipients having or not an AMR diagnosed within the 2 years after dn-iDSA-DQ detection; (B to E) Graft survival according to AMR or not combined with dn-iDSA-DQ active concentration (B), isolated dn-iDSA-DQ or multiple DSA at T0 (C), dn-iDSA-DQ MFI at T0 (D) and dn-iDSA-DQ persistence (E). AMR: antibody mediated rejection; dn-iDSA-DQ: de novo immunodominant donor-specific anti-HLA-DQ antibodies; SAFB: single antigen flow beads.

In multivariate analysis, the two variables included in the model which were independently associated with graft loss were AMR (HR 4.28, 95% CI 1.39 – 13.14, p = 0.01) and viral pulmonary infections (HR 3.31, 95% CI 1.24 – 8.83, p = 0.02) (Table 5). An active concentration ≥ 0.3 nM of dn-iDSA-DQ at T0, T1 and/or T2 (HR 2.39, 95% CI 0.90 – 6.33, p = 0.08) and multiple DSA (HR 2.74, 95% CI 0.88 – 8.51, p = 0.08) were included in the model but were not independently associated with graft loss in multivariate analysis, yet with a trend (Table 3). Persistent DSA was not included in the multivariate model.

**Table 3:** Factors within the 2 years after dn-iDSA-DQ development associated with graft loss.

|  | Univariate analysis |  |  | Multivariate analysis |  |  |
| --- | --- | --- | --- | --- | --- | --- |
|  | HR | 95% CI | p-value* | HR | 95% CI | p-value** |
| Quantifiable dn-iDSA-DQ active concentration within the 2 years after their detection | 2.84 | 1.10 – 7.30 | **0.03** | 2.39 | 0.90 – 6.33 | 0.08 |
| Multiple DSA at dn-iDSA-DQ detection | 2.55 | 0.84 – 7.71 | 0.10 | 2.74 | 0.88 – 8.51 | 0.08 |
| DSA persistence | 4.00 | 1.16 – 13.79 | **0.03** | NI |  |  |
| dn-iDSA-DQ MFI (above or below median MFI) at their detection | 1.65 | 0.64 – 4.23 | 0.30 |  |  |  |
| Anastomotic complications | 0.46 | 0.11 – 1.99 | 0.30 |  |  |  |
| CMV status | 1.03 | 0.54 – 1.99 | 0.92 |  |  |  |
| Biopsy-proven acute cellular rejection | 0.74 | 0.28 – 1.97 | 0.55 |  |  |  |
| Antibody-mediated rejection (AMR) | 3.99 | 1.31-12.10 | **0.02** | **4.28** | **1.39 – 13.14** | **0.01** |
| Plasma exchange and/or rituximab | 2.91 | 1.13-7.50 | **0.03** | NI |  |  |
| Bacterial pulmonary infection | 1.83 | 0.71 – 4.70 | 0.21 |  |  |  |
| Fungal pulmonary infection | 1.50 | 0.56 – 4.01 | 0.42 |  |  |  |
| SARS-Cov2 pulmonary infection | ND*** |  |  |  |  |  |
| Other viral pulmonary infection | 3.62 | 1.36 – 9.60 | **0.01** | **3.31** | **1.24 – 8.83** | **0.02** |

CI: confidence interval; HR: hazard-ratio; ND: not determined; NI: variables not included in the multivariate model through backward exclusion;
\*Cox hazard regression and \*\*backward multivariate analysis including factors with p<0.2 in univariate analysis. The overall model fit significance level was p = 0.008. \*\*\*Not determined because of an insufficient number of events.

## Discussion

We report for the first time, to our knowledge, that the active concentration of dn-iDSA-DQ, measured with SPR in the serum of their first detection with SAFB, is associated with CLAD development after lung transplantation. Importantly, despite the retrospective design our work, we were able to study dn-iDSA-DQ concentration as early as possible in the natural history of allogeneic humoral response of these LTRs.

Another important finding is that, despite a limited correlation, dn-DSA-DQ active concentration cannot be predicted by SAFB MFI. This suggests that quantitative measurement of these DSA by SPR could bring unique additional information about their pathogenicity. This could be due to the fact that the SAFB MFI is likely a marker integrating multiple parameters: 1) the active concentration of the antibody, 2) its affinity, 3) the density of HLA molecules on the surface of the bead and 4) the existence of various interferences for IgG detection.

In addition, the active concentration of dn-iDSA-DQ wad associated with graft survival in univariate analysis, and was trending in multivariate analysis. The diagnosis of AMR, integrating the presence of DSA and pathology features evidenced on lung transplant biopsies, was the most powerful parameter associated with graft loss. The association of AMR with other DSA parameters such as SAFB MFI, the number of DSA, their active concentration and their persistence, evidenced worse prognosis in univariate analysis. Among the three parameters that would be available at the earliest time, active concentration and the number of DSA appeared to be the most valuable to guide clinical decision. Noteworthy, only 8 LTRs had an AMR diagnosed together with dn-iDSA-DQ detection, meaning that AMR was more often diagnosed after dn-iDSA-DQ development. One could propose to reinforce follow-up, seeking for AMR, or even to initiate AMR treatment when active concentration is quantifiable and/or multiple DSA are present at their first appearance.

Persistent DSA with SAFB are of worse prognosis in several studies (13,22–25) yet several weeks of follow-up are required to obtain this information, which could delay treatment in some LTRs who need it. In our cohort, each dn-iDSA-DQ which had an active concentration ≥ 0.3 nM at T0 persisted over time, which reinforce the potential clinical application of measuring dn-iDSA-DQ active concentration when they are first detected.

The SPR assay would be a complementary assay devoted to refining the characterization of DSA identified by SAFB. This could improve LTR’s clinical management. Noteworthy, the serum pre-treatment process takes only 3 to 4 hours. Then, CFCA takes about 3 hours for a sample, which means that SPR results can be provided within 1 to 2 working days. This is fully compatible with the clinical practice.

The SPR assay suffers from several drawbacks. The most important, in our opinion, is that a non-negligible proportion of LTRs followed in our centres could not be included in our study because the SPR anchor competed with their dn-iDSA-DQ. Further work is needed to generate alternative anchors recognizing different areas of HLA-DQ molecules, which would allow studying active concentration of dn-iDSA-DQ in all LTRs. Of note, SPR also allows to study DSA directed against class I HLA (33,35), yet we decided to focus on anti-DQ DSA which are the most frequent in lung transplantation and of worse prognosis. More than half of the LTRs in our cohort also had multiple DSA. Because of the retrospective design of our study, amount of serum was not sufficient to measure the active concentration of every DSA produced. The amount of serum required to be purified and then to be processed by SPR to measure active concentration is another drawback, which prevented the inclusion of numerous LTRs in this retrospective study. This drawback would not be of further importance in a prospective setting.

Importantly, in our setting, working on purified IgG was of outstanding importance for several reasons. The first one was to study IgG DSA only, which are considered as the most relevant DSA. Second, it is known that patients developing de novo IgG DSA are also generating IgM DSA, which would be especially the case at the first time point studied in our cohort. Because IgM DSA can interfere with IgG when using SAFB and SPR assays (18), working on purified IgG allowed us to obtain reliable results. Third, as previously described, working on purified IgG decreases non-specific binding of serum components on the SPR surface. This further increases the reliability of our method (34).

Noteworthy, our method also allows to determine anti-HLA-DQ antibodies affinity. However, to provide reliable results, the active concentration of the antibodies must be high enough, which was the case for a limited number of LTRs in our study (data not shown). Alternatively, in a prospective setting, it could be possible to purify a higher amount of serum to obtain a purified IgG sample with higher DSA concentration. However, we anticipate that this would be successful in only few cases.

Schneider *et al.* and Hug *et al.* recently used immobilization-free ligand binding technologies for in-solution analysis of anti-HLA antibodies to determine their concentration and their affinity directly in the original serum matrix (39,40). These methods determine the probability distribution of antibody concentration and interaction affinity at the same time, which unfortunately results in large confidence intervals for these quantitative parameters. Moreover, to obtain significant binding and thus reliable and concomitant affinity and concentration measurements, the antibody binding site concentration has to significantly exceed the dissociation equilibrium constant, which lies in the nanomolar range (33–35,39,40). Based on our results showing that active concentrations of dn-iDSA-DQ are rarely above 10 nM, we anticipate that alternative methods requiring concomitant affinity and concentration measurements would not provide the same accuracy as CFCA by SPR does. The method we use is only based on mass transport limitation of DSA, a parameter that is only dependent of the diffusion coefficient of IgG. Finally, Hug et al. showed that the presence of IgM DSA together with IgG could drastically alter flow-induced dispersion analysis results (39), which cannot happen in our method which uses purified IgG. Whether this matters for a clinical application would need further evaluation.

Altogether and thinking of the best diagnostic arsenal to be used in the future, we believe that additional and collaborative works are needed to determine the complementarity of the different methods allowing to measure DSA concentration. Regarding affinity determination and for both SPR and in-solution assays, the low active concentration of DSA is a limitation. This means that a direct determination of the affinity of all serum DSA will not be easily doable with current available methods for a clinical application.

In conclusion, our study shows that measuring the active concentration of dn-iDSA-DQ by SPR is a potential new biomarker for predicting CLAD-free survival and graft loss. This work paves the way for further prospective investigations on larger prospective cohorts to determine whether the clinical management of LTRs could benefit from this SPR-based methods or others.

## Data Availability

All data produced in the present study are available upon reasonable request to the authors.

## Acknowledgments

This work has benefited from the facilities and expertise of the Structural Biophysico-Chemistry plateform (BPCS) at IECB, CNRS UMS3033, Inserm US001, University of Bordeaux (http://www.iecb.ubordeaux.fr/index.php/fr/plateformestechnologiques), giving access to the Biacore T200 instrument that was acquired with the support of the *Conseil Régional d’Aquitaine*, the GIS-IBiSA and the *Cellule Hôtels à Projets* of the CNRS. We thank Lætitia Minder and Jean-Michel Blanc for technical assistance with the Biacore T200. We thank Thermofisher, One Lambda for providing the purified HLA-DQ antigens.

## Abbreviations

ACR: acute cellular rejection
AMR: antibody-mediated rejection
CFCA: calibration-free concentration assay
CLAD: chronic lung allograft dysfunction
dn-iDSA-DQ: *de novo* immunodominant anti-DQ DSA
DSA: donor specific antibodies
HLA: human leukocyte antigen
LTRs: lung transplant recipients
MFI: mean fluorescence intensity
SAFB: single antigen flow beads
SPR: surface plasmon resonance

