## Supplementary material for "Active concentration of de novo anti-HLA-DQ donor specific antibodies measured by surface plasmon resonance is associated with chronic lung allograft dysfunction": Suppl Figures

**Supplemental Figure 1: Study flow chart.**

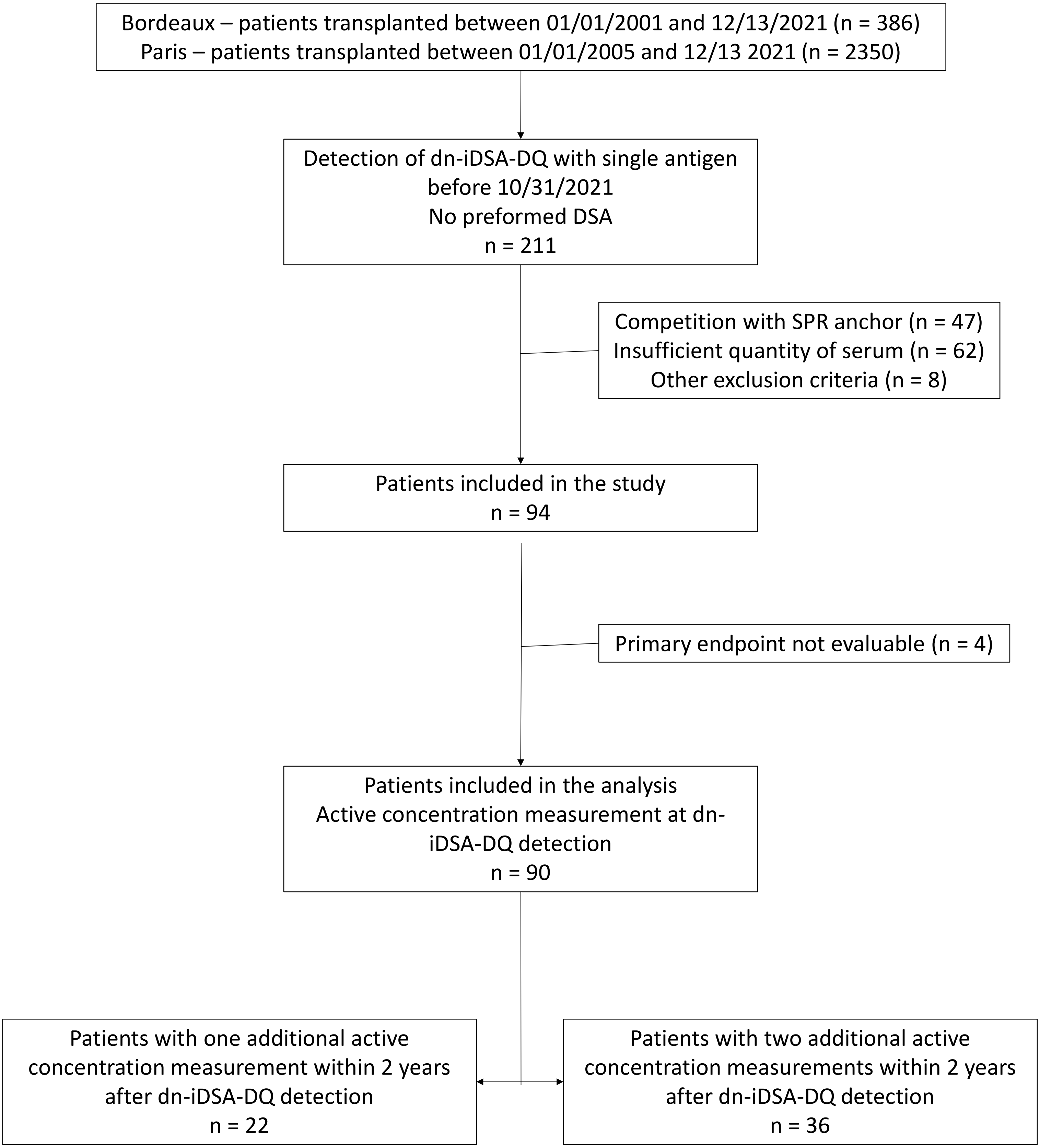

**Supplemental Figure 2: Principle of Anchor competition experiments**

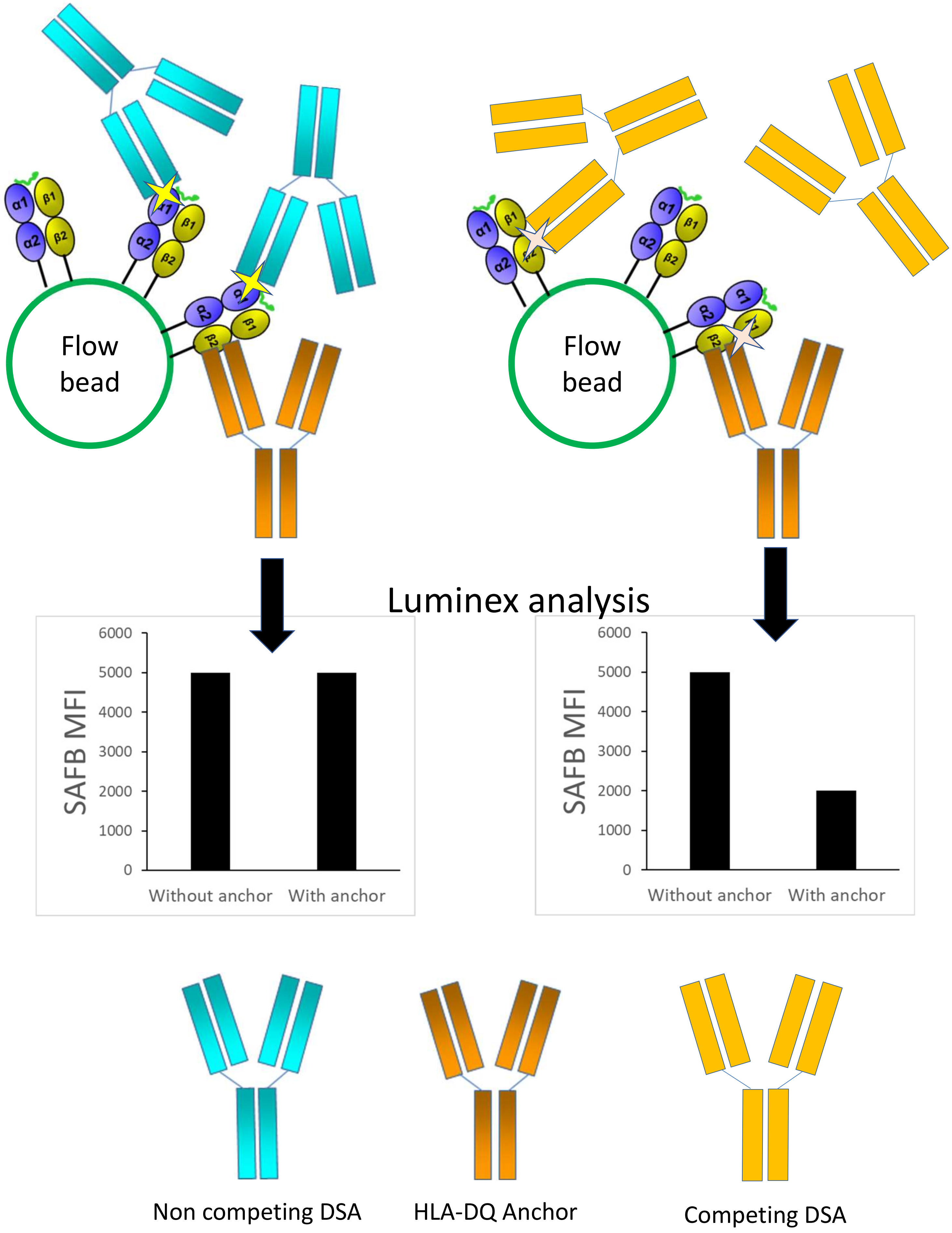

**Supplemental Figure 3: Workflow for active concentration measurement**

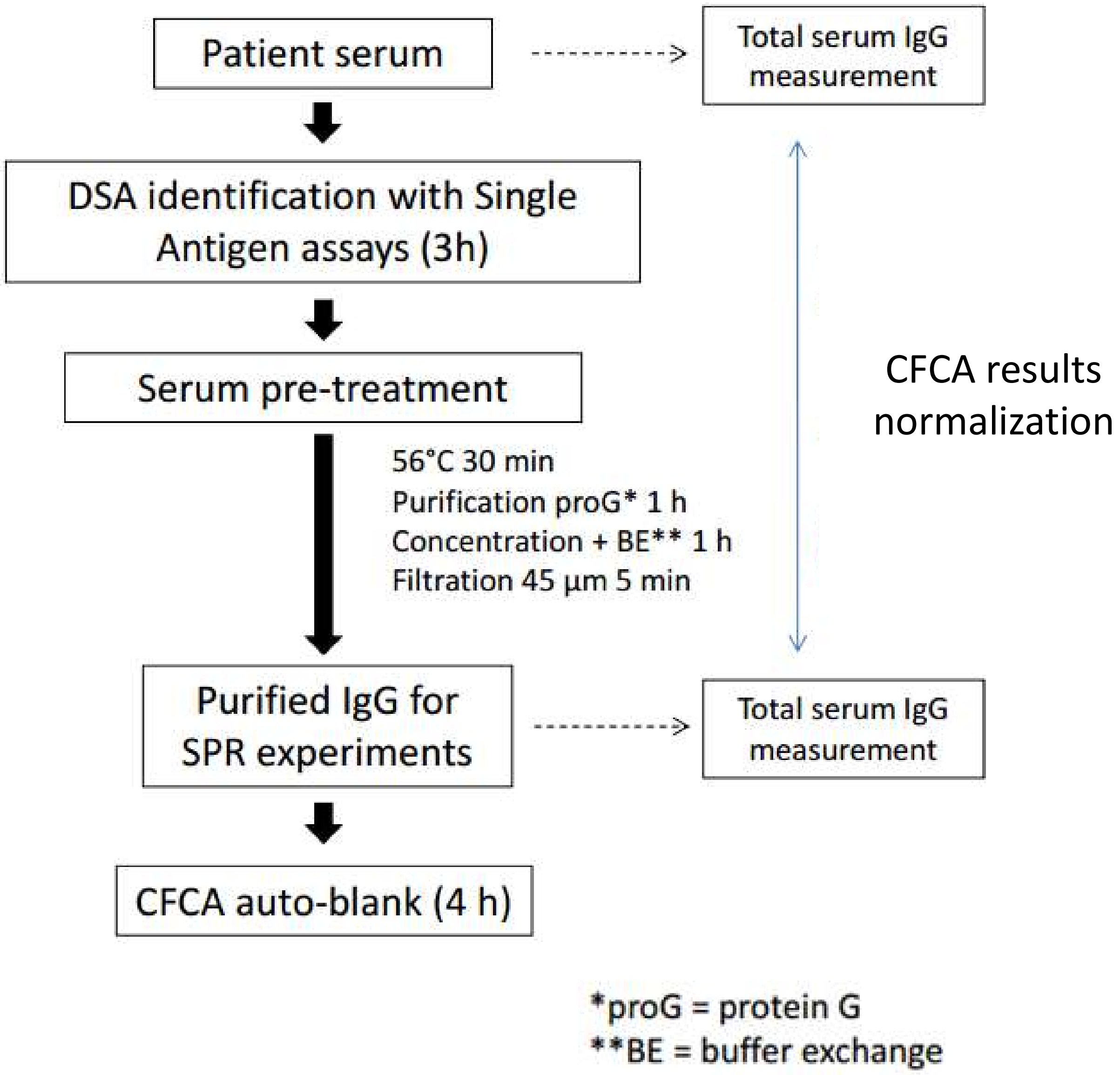
