## Supplementary material for "Active concentration of de novo anti-HLA-DQ donor specific antibodies measured by surface plasmon resonance is associated with chronic lung allograft dysfunction": Suppl Tables

**Supplemental Table 1 – Recipients’ characteristics before dn-iDSA-DQ development**

|  | **n = 90 recipients** |
| --- | --- |
| **Delay between transplant and DSA detection [median (1Q/3Q), months]** | 3 (1-10) |
| **Anastomotic complications (n, %)** | 14 (15.6%) |
| **Cancer (n, %)** | 4 (4.4%) |
| **Acute cellular rejection (n, %)** | 30 (33.3%) |
| **Bacterial pulmonary infection (yes/no)** | 67 (74.4%) |
| **Fungal pulmonary infection (n, %)** | 34 (37.8%) |
| **SARS-Cov2 pulmonary infection (n, %)** | 2 (2.2%) |
| **Other viral pulmonary infection (n, %)** | 18 (20.0%) |

**Supplemental Table 2: Factors from transplantation to dn-iDSA-DQ detection associated with CLAD up to 5 years after DSA detection**

|  | **Univariate analysis** | | |  | **Multivariate analysis** | | |
| --- | --- | --- | --- | --- | --- | --- | --- |
|  | **HR** | **95% CI** | **p-value*** |  | **HR** | **95% CI** | **p-value**** |
| **Quantifiable dn-iDSA-DQ active concentration at their detection** | 1.79 | 1.04 – 3.10 | **0.04** |  | **2.00** | **1.13 – 3.56** | **0.02** |
| **Multiple DSA at dn-iDSA-DQ detection** | 2.49 | 1.37 – 4.53 | **0.003** |  | **2.85** | **1.55 – 5.23** | **0.0008** |
| **dn-iDSA-DQ MFI (above or below median MFI) at their detection** | 1.59 | 0.92 – 2.73 | 0.10 |  | NI |  |  |
| **Anastomotic complications** | 0.86 | 0.41 – 1.82 | 0.70 |  |  |  |  |
| **CMV status** | 1.13 | 0.78 – 1.63 | 0.53 |  |  |  |  |
| **Biopsy-proven acute cellular rejection** | 1.87 | 1.09 – 3.21 | **0.02** |  | **2.22** | **1.26 – 3.92** | **0.006** |
| **Bacterial pulmonary infection** | 1.95 | 0.98 – 3.88 | 0.06 |  | 1.89 | 0.92 – 3.89 | 0.09 |
| **Fungal pulmonary infection** | 1.89 | 1.10 – 3.23 | **0.02** |  | 1.50 | 0.85 – 2.64 | 0.16 |
| **SARS-Cov2 pulmonary infection** | ND*** |  |  |  |  |  |  |
| **Other viral pulmonary infection** | 1.48 | 0.79 – 2.76 | 0.22 |  |  |  |  |

CI: confidence interval; HR: hazard-ratio; ND: not determined; NI: variables not included in the multivariate model through backward exclusion; *Cox hazard regression and **backward multivariate analysis including factors with p<0.2 in univariate analysis. The overall model fit significance level was p < 0.0001. ***Not determined because of an insufficient number of events.

**Supplemental Table 3 – Association between DSA characteristics and AMR**

|  |  | **AMR** | | **Fisher’s exact test (p=)** |
| --- | --- | --- | --- | --- |
|  |  | **no** | **yes** |  |
| **Quantifiable dn-iDSA-DQ active concentration at their detection** | **no** | 31 | 28 | 0.38 |
|  | **yes** | 13 | 18 |  |
| **Quantifiable dn-iDSA-DQ active concentration within the 2 years after their detection** | **no** | 28 | 26 | 0.52 |
|  | **yes** | 16 | 20 |  |
| **Multiple DSA at dn-iDSA-DQ detection** | **no** | 25 | 20 | 0.29 |
|  | **yes** | 19 | 26 |  |
| **dn-iDSA-DQ MFI (above or below median MFI) at their detection** | **no** | 17 | 20 | 0.67 |
|  | **yes** | 27 | 26 |  |
| **DSA persistence** | **no** | 18 | 16 | 0.52 |
|  | **yes** | 25 | 30 |  |

AMR: antibody-mediated rejection
